# Snakebite epidemiology and health-seeking behaviour in fringe communities of the Kakum Conservation Area in Ghana

**DOI:** 10.1101/2025.06.26.25330342

**Authors:** Derek Anamaale Tuoyire, Linus Baatiema, Robert Peter Biney, Justus Precious Deikumah, John Koku Awoonor-Williams, Mawuli Kotope Gyakobo

## Abstract

**Background:** Snakebite envenomation (SE) remains a significant yet underreported public health challenge, especially in rural and forest-fringe communities in sub-Saharan Africa. Despite its recognition by the World Health Organization (WHO) as a Neglected Tropical Disease (NTD), epidemiological data on SE remain scarce in Ghana, particularly in communities fringing protected forest areas such as the Kakum Conservation Area (KCA).

**Objective:** This study aimed to quantify and characterize the epidemiology of snakebite envenomation and explore the health-seeking behaviours among residents in the fringe communities of the KCA in the Central Region of Ghana.

**Methods:** A cross-sectional household survey was conducted between April and May 2024 in 18 communities fringing the KCA in Ghana’s Central Region. A two-stage sampling approach was employed to enumerate a total of 1,445 households for snakebite victims. The survey instrument was programmed into KoBoCollect software and administered to snakebite victim of household heads using the one-on-one interview method. Mainly descriptive statistical analytical analyses were conducted using STATA version 14.0.

**Results:** In this study, the lifetime number of snakebites reported was 394 (4.6%), while 164 (1.9%) and 41 (0.5%) snakebites reported in the last 5 years and one (1) year preceding the survey, respectively. The average age of a snakebite victim was 44 years. Snakebite victims were typically female (54%), married (59%), and mainly engaged in farming (73%) for livelihood. Most snakebites occurred during the morning (42%), in the farm field/forest (66%) and along the victims’ lower limbs (88%). Over seven-in-ten victims sought first aid, with tourniquet (35%) mostly applied. Majority (69%) of victims sought treatment in a healthcare facility, mostly in Community-based Health Planning and Services (CHPS) compounds (32%).

**Conclusion:** Snakebites constitute a major public health concern in communities fringing the KCA, especially for those whose livelihoods revolve around agricultural activities. There is the need to establish and strengthen collaborative efforts among primary health authorities, forest conservation managers, and community leaders in the design and implementation of effective interventions to avert snakebite incidents and improve outcomes for victims.

**Author summary:** Snakebite envenomation is a serious yet underreported public health issue particularly in rural forest-fringe communities in Africa. Despite being reclassified by the World Health Organization as a Neglected Tropical Disease, snakebite remains poorly documented in Ghana, especially in communities bordering protected forests where human-snake encounters are more likely. We surveyed over 1,400 households in 18 communities surrounding the Kakum Conservation Area in southern Ghana to understand how common snakebites are, the kind of people affected, and how they seek treatment after snakebites. We found a relatively high number of snakebites, mostly affecting adult farmers while working on their farms or in nearby forests. Bites typically occurred on the legs and were often followed by symptoms such as swelling and bleeding. While over two-thirds of victims received some form of formal medical care, some continued to seek traditional remedies even after visiting a health facility.

Our findings suggest snakebite is a persistent health threat in rural forest-fringe communities and that health-seeking behaviour after snakebite is shaped by a combination of accessibility, cultural beliefs, and resource limitations. Therefore, strengthening rural health systems and improving community education at the local level are crucial steps to reduce the burden of snakebite in Ghana and similar settings.

## Background

Snakebite envenomation (SE) has increasingly become a public health concern, warranting its re-inclusion on the list of Neglected Tropical Diseases (NTDs) by World Health Organization (WHO) in 2017 [1, 2]. Although the actual number of snakebites and envenomation remain unknown, between 4.5 and 5.4 million people are estimated to experience snakebites annually across the world, accounting about 400,000 disabilities with mortality ranging from 81,000 to 138,000 [3-5]. Given that human-snakebite encounter typically occur during field related agricultural activities where there are high grass or forests among other factors, snakebites disproportionately affect those residing in rural communities in low- and middle-income countries in Asia, Latin America and Africa [6, 7]. In sub-Saharan Africa, snakebite envenomation afflicts an estimated 90,000 to 400,000 people annually, resulting in 3,500 to 32,000 deaths among victims [8-10].

Snakebite envenomation appears to be associated with livelihood activities given that victims are largely reported to be young and economically active; who are involved in subsistence farming, with bite sites often located on their extremities [11]. Given their underprivileged backgrounds, majority of victims are usually unable to seek prompt treatment in appropriate healthcare facilities, due in part to long distance and associated healthcare costs, poor knowledge and skepticism about the effectiveness of allopathic medicine in treating snakebite envenomation. As such victims often resort to traditional healers who offer remedies including use of tourniquets, incision at the site of bite, application of topical and emetic medicinal herbal concoctions or application of the black stone [12, 13]. These remedies may not be effective and could compound the complications of SE. This propels a mutually reinforcing vicious cycle of poverty, livelihood activities, snakebite envenomation and ineffective health seeking behaviours resulting in dire outcomes for victims. Survivors, may suffer physical consequences such as life-long disabilities including amputations [14, 15]. The psychological trauma, limitations on work and overall quality of life of snakebite victims cannot be overemphasized.

As in other regions across the globe, there is paucity of research and epidemiological data on the burden of snakebite envenomation in Ghana. Available studies vary in their reporting of the burden of snakebite envenoming. For instance, in northern Ghana, Musah et al. [11] estimated the prevalence of snakebite to be approximately 6%, with case fatality rate of 3% [11], while Yakubu et al. [16] reported about 86 envenomings, with an annual mortality estimated at 24/100,000. Another study Punguyire et al [17] found a snakebite incidence of 92/100,000 in the Brong-Ahafo region. A more recent review based on data from the District Health Information and Management Systems (DHIMS) in the Oti region of Ghana between 2014 and 2018 reported total of 2,973 cases of snakebite, with 5-year average incidence of 24/100,000.

While these studies provide some useful epidemiological insights into snakebite envenomation in Ghana, there is a dearth of similar studies in communities located within the fringes of forested areas, which typically serve as habitat for variety of distinct snake fauna and present unique human-snake interactions. Inhabitants of such communities are particularly more vulnerable to snakebite envenomation, as they tend to live in poor housing conditions within the frontiers of natural snake habitats, while engaged in subsistence agricultural activities and often several hours away from the nearest functional healthcare facility [18, 19].

This study sought to quantify and characterise snakebite envenomation in the fringe communities of the Kakum Conservation Area (KCA) in the Central region of Ghana. KCA is a protected area of moist semi-deciduous forest which forms part of the Upper Guinean Forest biodiversity hotspot of West Africa [20]. Cognizant of the WHO’s recognition of the lack of epidemiological awareness of snakebite envenomation and commitment to halving deaths and disabilities from snakebites by 2030 [6], this study would provide useful insights and contribute to the body of knowledge.

## Methods

### Study setting and design

The study was conducted in communities fringing the KCA, located within the Central region of Ghana. The KCA is located at 5.4167N and -1.3167W, covering an estimated 360 km^2^ comprising of two contiguous reserves – the Kakum National Park (210 km^2^) and the adjacent Assin Attandanso Resource Reserve (150 km^2^). This combined protected area forms part of the Upper Guinean Forest biodiversity hotspot of West Africa [20] and is characterized by three principal forest types: moist forest, edaphic forest, and boval forest [21, 22].The moist forests can be divided into two subgroups: Lophira alata - Triplochiton scleroxylon forest and Celtis zenkeri – T. scleroxylon forest. The edaphic forests are comprised of swamp and riverine forest. The population of the area is about 119,359 with about 64 fringe communities.

Given that the KCA is surrounded by farming communities in a 360^°^ fashion, a two-staged sampling cross-sectional household survey approach was employed. In the first stage the KCA was zoned according to the eight (8) main geo-cardinal points (North, South, East, West, North-East, North-West, South-East and South-West) surrounding the forest, based on which the simple random sampling techniques was used to select at least two (2) fringe communities to represent each geo-cardinal zone (**Figure 1**). The next level of selection involved a door-to-door enumeration of all households within each selected community. The primary respondent to ascertain the presence or otherwise of a snakebite victim(s) at the household level was the head of household. Consenting/assenting snakebite victims identified within households were then subsequently interviewed.

**Fig 1:**
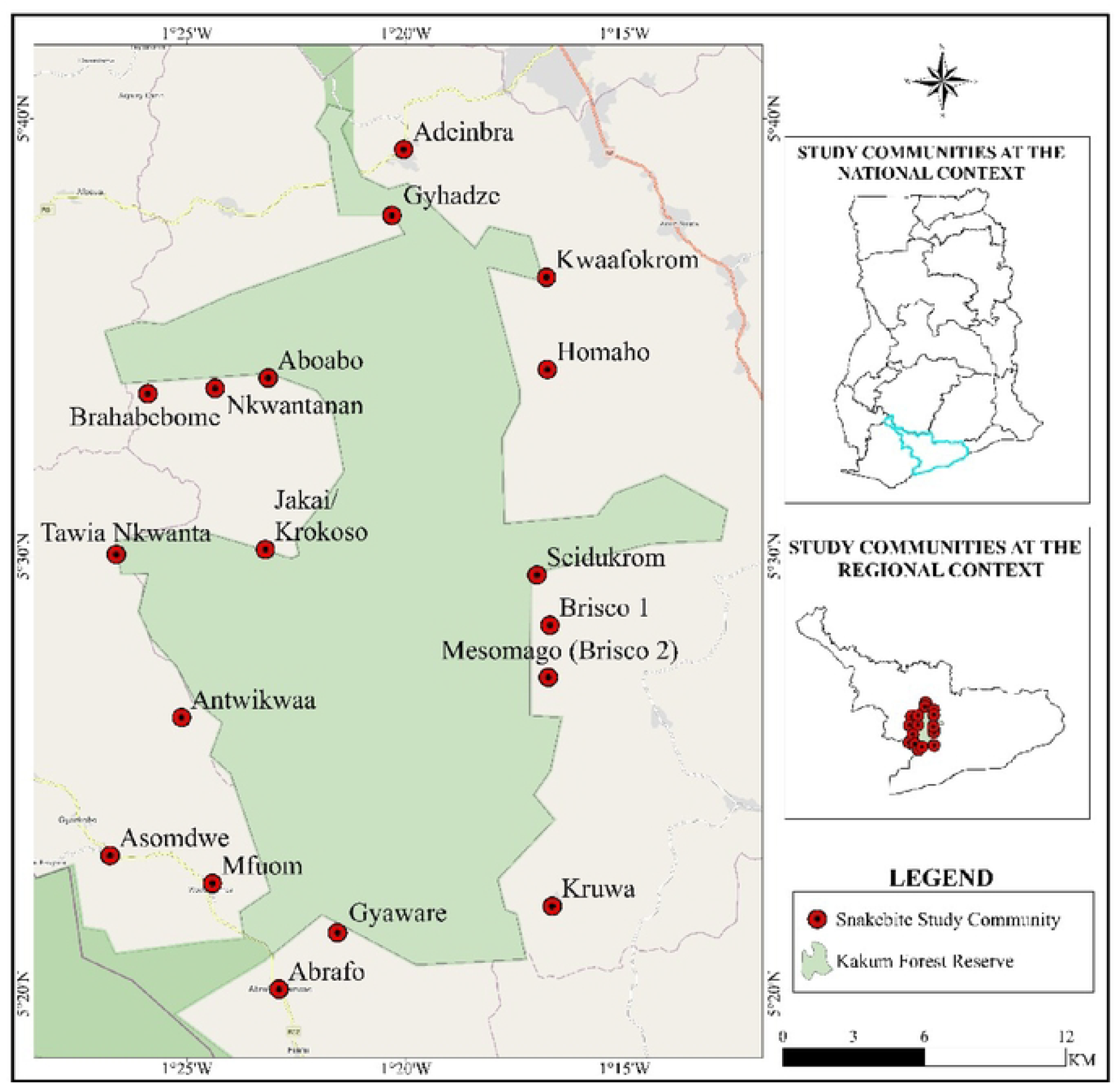
Geo-cardinal points and selected communities in the Kakum Conservation Area (KCA)

### Study population and sampling

The study targeted two categories of participants in the peripheral communities within the KCA forest enclave. The first consisted of snakebite victims who were bitten in the last five (5) years prior to the commencement of the study, while the other category consisted of traditional healers with experience of managing snakebite victims. A two-staged cross-sectional survey approach was employed in the selection of participants in the first phase (quantitative) of data collection. Sample communities were selected to represent each geo-cardinal points (North, South, East, West, North-East, North-West, South-East and South-West) surrounding the forest, based on which one (1) or more communities were selected to represent each geo-cardinal zone using simple random sampling and Arc-GIS tool. A total number of 18 communities (**See Figure. 1**) were selected from which 1,445 households were enumerated.

### Data collection

Data collection for the study took place between April and May 2024, involving two (2) mobile data collection teams each consisting of four (4) enumerators and two (2) supervisors and a local park ranger as a guide and liaison. All team members participated in two-day training tailored to the purposes of the project. The data collection involved the door-to-door enumeration of household heads and snakebite victims using electronic tablets with questions programmed using the KoBoCollect software [23, 24]. The data collection instrument had a section for interviewing the head of household and another section for snakebite victims linked with their household related information which could only be activated once a snakebite victim was identified within a household. The data collection instrument mainly covered household environment and socio-demographic information, snakebite experience, snake and snakebite characteristics, symptoms and treatment trajectories, as well as outcomes of the snakebite. Interviews were conducted in either English Language or the local language (Fante/Twi), depending on the preference of the respondent. All enumerators were proficient in both English and Fante/Twi languages.

### Data analysis

The data cleaning and analysis were conducted using STATA 14.0 (StataCorp LLC). Mainly descriptive techniques were employed in the data analysis. The prevalence of snakebite was estimated with reference to the last five (5) years preceding the survey. This was calculated using a count of each household size as the denominator and expressed with 95% confidence intervals (CI). This was followed by the use of frequency distributions (proportions) to describe and characterize the experiences and treatment trajectory of snakebite victims.

### Ethical approval

Ethical approval, reference UCCIRB/EXT/2023/22 for the study was obtained from the University of Cape Coast Institutional Review Board. Prior to data collection, informed consent was sought and obtained from all participants. For participants who were minors or incapacitated due to snakebite-related complications, consent was obtained from a parent, guardian, or responsible adult household member. Participation in the study was entirely voluntary, and respondents were informed of their right to withdraw at any stage without a penalty. All data were anonymized to ensure confidentiality and were securely stored to prevent unauthorized access. Interviews were conducted in the participants’ preferred language (English, Fante/Twi) to facilitate comprehension and comfort. The study adhered to the ethical principles outlined in the Declaration of Helsinki.

## Results

### Snakebite prevalence and outcomes

Out of the 1,445 households (8,518 inhabitants) enrolled in the study. The lifetime number of snakebites reported was 394 (4.6%), with 164 (1.9%) and 41 (0.5%) snakebites in the last 5-years and 1-year preceding the survey, respectively (**Table 1**). In terms of community specific prevalence of snakebites (**Figure 2**), there were no reports of snakebite in Gyaware in the last 5 years preceding the survey. However, Brisco-1 and Jakai/Krokoso reported a similarly higher annual prevalence of snakebites (2.3%); while Gyahadze had an even higher (4.6%) 5-year prevalence of snakebites compared with the other communities fringing KCA. In total, six (6) deaths from snakebites were reported, with 3 in the 5-year period before the study and one (1) in the last year prior to the study.

**Figure 2:**
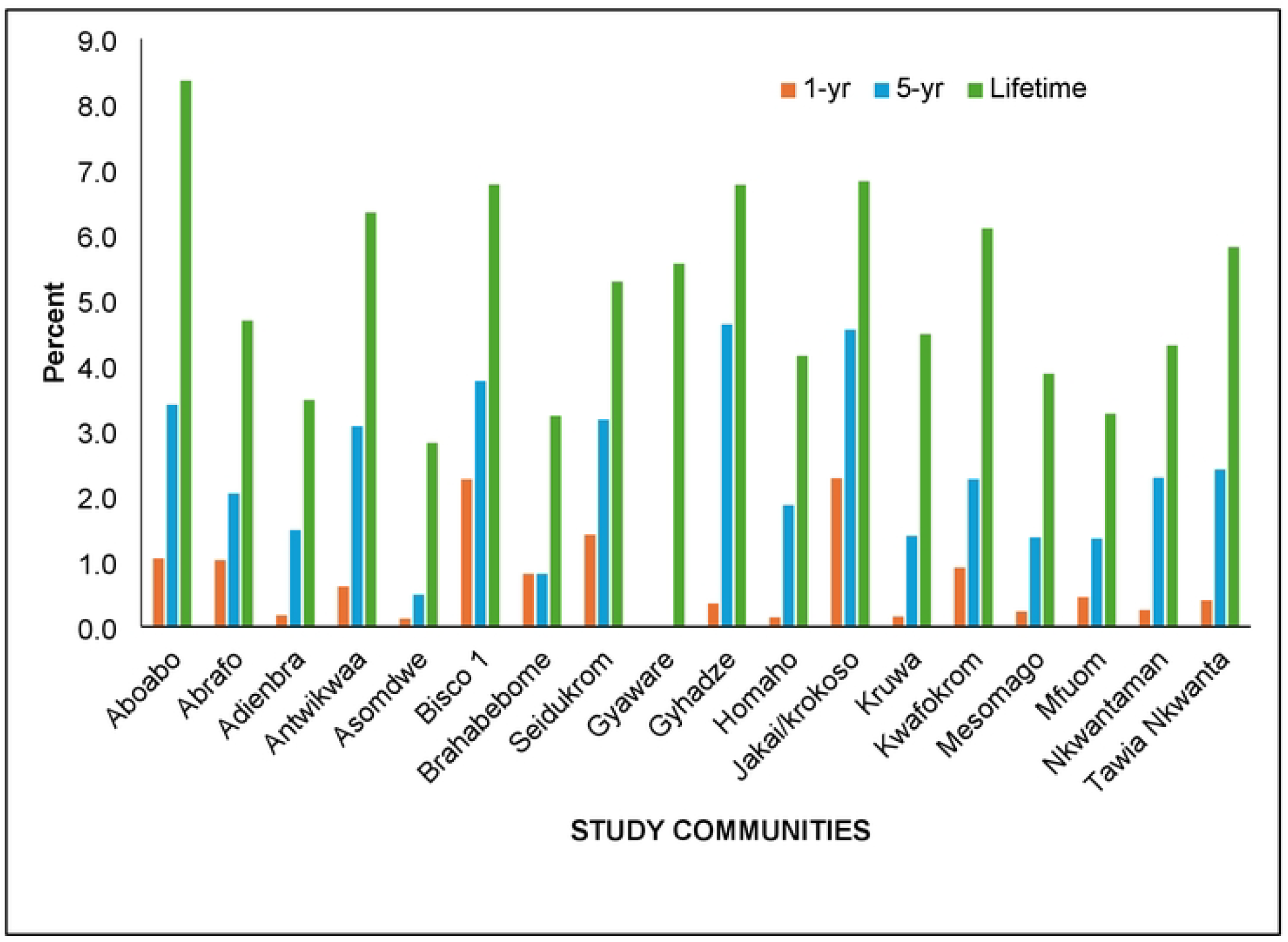
Community specific prevalence of snakebites in the KCA.

### Socio-demographic characteristics of snakebite victims

The mean age of a typical snakebite victim in the communities fringing KCA was 44 years (±18.6). There was a preponderance of females (54%) and most victims had JHS/JSS/Middle School (47%) education. About 59% of victims were married, with seven out of ten engaged in farming as their main occupation. For majority of victims (72%), the Community-based Health Planning and Services (CHPS) was the main facility by which they accessed orthodox healthcare. Only about 5% of the victims utilized unorthodox healthcare services (**Table 2**).

### Snakebite characteristics

Attributes of the perpetrating snake and other characteristics relating to the event of the snakebite are presented in **Table 3**. Most snakes were reported to be brown (26%) in colour and about 3-5 meters long (44%). More than eight-in-ten of victims were bitten along their lower limbs (leg-foot-knee). About 41% of snakebites were reported to have occurred in the morning, with over six out of ten of such bites taking place in the farm field or forest. Pain with bleeding (29%) was the most commonly reported symptom following the snakebite, followed by pain with swelling (26%).

### Snakebite treatment trajectory

The treatment trajectory following a snakebite is presented in **Table 4**. About 75% of victims indicated that they sought first aid after the snakebite incident, with over a third (35%) of them resorting to tourniquet as first aid. About two-thirds of victims sought care beyond the first aid, mostly from CHPS compounds (32%) while about a quarter of them sought care from herbalists. For about two-thirds of the victims, both the wait-time and journey between the snakebite incident and care-seeking was mostly less than an hour. Trekking on foot (40%) was the most common means of transports from the site of the snakebite incident to the place of care, while about a fifth of victims traveled by a taxi. Once at the place of care, a wide variety of treatment modalities were applied, however, the application of herbal remedies (21%) seemed to be the singular most common treatment. About a quarter of those who sought care in orthodox healthcare facilities later sought complementary care from traditional healers.

## Discussion

This study investigated the magnitude and characteristics of snakebite, snakebite envenomation and complications in the fringe communities of the KCA in the Central Region of Ghana. We found a high prevalence of snakebites in the rural communities fringing the KCA, which is consistent with existing literature indicating that rural dwellers, particularly those living close to forest reserves and wildlife parks, are more exposed to snake-human interactions [25, 26].The proximity of the KCA to these fringe communities likely increases the risk due to frequent encroachment into natural habitats for farming, fuel wood collection, and other livelihood activities. Other studies in sub-Saharan Africa and rural Asia affirm this high prevalence, noting that poor housing conditions, low awareness, and limited protective clothing contribute to increased exposure [27]. However, some recent interventions, such as awareness campaigns and habitat fencing in other parts of Ghana, have shown reductions in snakebite incidence [11], suggesting that targeted preventive efforts could be effective in KCA as well.

The study identified agricultural workers as the most affected group, reaffirming findings by Sharma et al. [28] and Arias-Rodríguez and Gutiérrez [29], who emphasized the vulnerability of farmers due to their frequent interactions with snake-prone environments. Farming activities, especially those involving weeding and harvesting, inadvertently increase the chances of stepping on or disturbing snakes. This finding aligns with Ghanaian studies in the Northern Region, where over 70% of snakebite victims were crop farmers [11]. These findings call for occupation-specific protective measures, including the distribution of boots, gloves, and community training on identifying high-risk zones during peak farming seasons.

The study revealed a relative preponderance of brown coloured snakes (26.9%); and snake length <2m amounted to 27.2%. These findings are consistent with the description of the viperidae, saw-scaled viper (*Echis ocellatus*) a finding corroborated by [16] who found this species as the most common cause of snakebite envenomation in Ghana. The saw-scaled viper is terrestrial and likely to strike the leg-foot-knee during an attack or bite (87.6%). This analysis may suggest why most bites occurred on the farm and forest areas. The study also revealed site/bleeding (29.2%) and site pain/swelling (25.9%) as the highest occurring symptoms and these are consistent with haemorrhagic and cytotoxic toxins which is an expressed venom of the viperidae Echis ocellatus [30]. Meanwhile, snake colours green and black and lengths >3m were also identified suggesting bites from elapids like the Mambas (*Dendroaspis spp*) and Cobras (*Naja spp*) respectively but without concomitant major neurologic symptoms reminiscent of dry bites. The recognition of multicoloured-blotched snakes adds another layer to the variety of snakes thus providing compelling reason for clinicians to review victims for clinical features before choosing antivenoms for treatment [30].

Most snakebites occurred during the morning and afternoon hours. This temporal pattern corresponds with farming hours, confirming that exposure increases with fieldwork activities. Studies by Warrell [31] and Iliyasu et al. [32] similarly found peak snakebite times to coincide with early and mid-day farming sessions. However, a contrasting study in Nepal reported evening and night bites as most common due to indoor incidents or bites while sleeping [33]. The variation likely stems from different snake species and community habits consistent with the mixed snake typology and varying occupational types expressed in this study but with skewness towards diurnal habits of the snakes and predominantly farming as an occupation. In KCA, where the population is active in the bush during daylight, this daytime pattern makes logical sense. This suggests the need for public education efforts to incorporate time-sensitive cautionary measures and training, particularly targeting the early and mid-day field periods.

The study also highlights the importance of the CHPS compounds as the primary access points in the continuum for care for snakebite envenomation. This reflects Ghana’s policy on decentralizing primary healthcare services and is consistent with findings from Sumah et al. [34] that CHPS compounds play a pivotal role in rural emergency response. Although our findings suggest that CHPS facilities are the preferred point of care for snakebites, perhaps due to proximity and perceived accessibility we are unable to ascertain from this study the specific form of treatment victims received. However, over a third (32%) of snakebite victims in this study received some form of non-orthodox treatment including herbs on wound, black stone and snake bile. This could plausibly reflect the lack of anti-venom and skilled personnel in the CHPS facilities visited, as prior studies [35, 36] have noted. Perhaps this is one area worth exploring further in subsequent studies. This may imply that even with limited resources, CHPS compounds are trusted entry points into the health system. Thus, strengthening CHPS compounds with snakebite treatment kits and referral systems could significantly improve outcomes and reduce complications or fatalities in these communities.

The study observed minimal delays in snakebite related health-seeking behaviour, which diverges from findings in other rural African settings where victims often resort to traditional healers before visiting formal health centers [37-39]. The reduced delays may be attributed to improved community awareness, health education campaigns, or previous negative experiences with traditional treatment outcomes. This finding contrasts with a study in rural Nigeria where up to 66% of victims first sought herbal treatment [40]. However, in a study in Northern Ghana, similar improvements in health-seeking behaviour were recorded in communities that had sustained public health campaigns on snakebite [41], supporting the idea that awareness and trust in the formal health system are growing. This is a promising development which could be leveraged further by integrating community volunteers and local champions to serve as first responders or referral agents during emergencies.

### Study Strengths and Limitations

This study provides valuable insight into snakebite trends and health-seeking behaviours in fringe communities of the Kakum Conservation Area. The key strengths of this study include the use of community-level data to situate our findings to local contexts, and its relevance for primary health care planning and delivery. The findings are also timely and useful for informing public health strategies aimed at reducing snakebite-related morbidity and mortality.

However, the study is limited by design structure. Its cross-sectional design limits causal interpretations, and reliance on self-reported data may introduce recall bias. Additionally, the findings are context-specific and may not be generalizable to other regions. The absence of clinical verification also means the severity and outcomes of snakebites could not be independently confirmed.

### Implication for Policy

The findings of this study have several key implications for health policy in Ghana. First, there is a critical need to strengthen CHPS compounds particularly in the study area by equipping them with essential snakebite management resources, such as anti-venom, snakebite kits, and trained personnel, particularly in high-risk rural areas. Policies should also integrate snakebite prevention and treatment into national health and occupational safety programmes, especially for agricultural workers. The demonstrated effectiveness of community health education in promoting prompt care-seeking behavior suggests that continuous public sensitization campaigns for snakebites should be supported and expanded. Moreover, ensuring the distribution of protective gear through agricultural extension services can reduce occupational exposure. Improved data collection and surveillance mechanisms such as incorporating comprehensive snakebite reports into the District Health Information Management System (DHIMS) are necessary for evidence-based planning. Finally, policy frameworks should promote collaboration between the health sector, forestry and wildlife authorities, agricultural sector and local governance structures to develop coordinated strategies for snakebite prevention, habitat management, and emergency response.

## Conclusion

Snakebites constitute a major public health concern in communities fringing forested conservation areas, especially for people whose livelihood activities revolve around agricultural activities. Most bites occur during morning and afternoon hours when farming activities are at their peak. Overall, the study underscores the need to establish and strengthen collaborative efforts among primary health authorities, forest conservation managers, and community leaders in the design and implementation of effective interventions to avert snakebite incidents and improve outcomes for victims.

## Data Availability

Available on request from DRIC, UCC

## Acknowledgement

The authors gratefully acknowledge the financial support provided by the Directorate of Research and Innovation and Consultancy of the University of Cape Coast (DRIC) for this research. We extend our sincere appreciation to Mr. Alex Agyei, the manager of the Kakum Conservation Area, for granting us permission and forest guards to facilitate navigation within the conservation area to conduct this study. Special thanks are due to Dr. Prince Anku and Sandra Akua Agyemang, who expertly managed the enumerators and logistics for this project. We also thank the dedicated enumerators for their hard work and commitment to data collection, and to Mr. Samuel Asiamah Nyave for helping with mapping. Finally, we express our gratitude to the anonymous reviewers for their valuable feedback and to the members of the study fringe communities near the Kakum Conservation Area for their participation and cooperation.

## Author contribution

**Conceptualization:** Derek Anamaale Tuoyire, Robert Peter Biney, Justus Precious Deikumah, Mawuli Kotope Gyakobo

**Data curation:** Derek Anamaale Tuoyire, Justus Precious Deikumah, Mawuli Kotope Gyakobo

**Formal analysis:** Derek Anamaale Tuoyire, Justus Precious Deikumah, Mawuli Kotope Gyakobo

**Funding acquisition:** Robert Peter Biney, Justus Precious Deikumah, Mawuli Kotope Gyakobo

**Investigation:** Derek Anamaale Tuoyire, Robert Peter Biney, Justus Precious Deikumah, John Koku Awoonor-Williams, Mawuli Kotope Gyakobo

**Methodology:** Derek Anamaale Tuoyire, Robert Peter Biney, Justus Precious Deikumah, Mawuli Kotope Gyakobo

**Project administration:** Robert Peter Biney, Justus Precious Deikumah, Mawuli Kotope Gyakobo

**Resources:** Derek Anamaale Tuoyire, Justus Precious Deikumah, Mawuli Kotope Gyakobo

**Software:** Derek Anamaale Tuoyire, Linus Baatiema,

**Supervision:** Derek Anamaale Tuoyire, Justus Precious Deikumah, Mawuli Kotope Gyakobo

**Validation:** John Koku Awoonor-Williams, Mawuli Kotope Gyakobo

**Visualization:** Justus Precious Deikumah

**Writing – original draft:** Derek Anamaale Tuoyire, Linus Baatiema

**Writing – review & editing:** Derek Anamaale Tuoyire, Linus Baatiema, Robert Peter Biney, Justus Precious Deikumah, John Koku Awoonor-Williams, Mawuli Kotope Gyakobo

